# Prevalence and clinical profile of SARS-CoV-2 infection among farmworkers in Monterey County, California: June-November, 2020

**DOI:** 10.1101/2020.12.27.20248894

**Authors:** Joseph A. Lewnard, Ana M. Mora, Oguchi Nkwocha, Katherine Kogut, Stephen A. Rauch, Norma Morga, Samantha Hernandez, Marcus P. Wong, Karen Huen, Kristin Andrejko, Nicholas P. Jewell, Kimberly L. Parra, Nina Holland, Eva Harris, Maximiliano Cuevas, Brenda Eskenazi, on behalf of the CHAMACOS-Project-19 Study Team

**Author notes:** Address for correspondence: Joseph A. Lewnard, 2121 Berkeley Way, Room 5410, Berkeley, California 94720. These authors contributed equally to the work. Members of the CHAMACOS-Project-19 Study Team are listed in the **Acknowledgements** section of the manuscript.

## Abstract

As essential personnel, United States farmworkers have continued working in-person throughout the COVID-19 pandemic. We undertook prospective surveillance of SARS-CoV-2 infection and antibody prevalence among farmworkers in California’s Salinas Valley from 15 June to 30 November, 2020. Over this period, we observed 22.1% (1514/6864) positivity for current SARS-CoV-2 by nucleic acid detection among farmworkers tested at federally-qualified migrant and community health clinics, as compared to 17.2% (1255/7305) among other adults from the same communities (risk ratio, 1.29; 95% confidence interval, 1.20-1.37). In a nested study enrolling 1,115 farmworkers, prevalence of current infection was 27.7% among farmworkers reporting ≥1 potential COVID-19 symptom, and 7.2% among farmworkers without symptoms (adjusted odds ratio 4.17; 2.86-6.09). Prevalence of anti-SARS-CoV-2 IgG antibodies increased from 10.5% (6.0-18.4%) between 16 July-31 August to 21.2% (16.6-27.4%) between 1-30 November. The high observed prevalence of infection among farmworkers underscores the need for vaccination and other preventive interventions.

## INTRODUCTION

In response to the ongoing coronavirus disease 2019 (COVID-19) pandemic, the United States and other countries have implemented broad interventions aiming to mitigate community transmission of severe acute respiratory syndrome coronavirus 2 (SARS-CoV-2) (*1*). Workers in food production and other industries deemed essential to continuity of public health and safety have continued in-person work (*2*). While COVID-19 outbreaks have been reported among various essential workforce groups, including employees in food processing facilities (*3, 4*), to date no studies have prospectively assessed risk of infection among essential workers involved in food production.

Agriculture and related food production industries comprise one of the lowest-paid sectors of the US economy, with 29% of full-time workers earning an annual income below $12,760 as individuals or $26,200 for a family of four (*5*). Agriculture in particular draws on a predominantly Latino immigrant workforce (*6*), who in comparison to their US-born counterparts work longer hours, receive lower wages, and experience higher levels of household poverty (*7*). An estimated 54% of immigrant farmworkers are undocumented, and thus have reduced access to federal benefits under the Coronavirus Aid, Relief, and Economic Security Act (*8*). These circumstances have compounded pre-existing legal and economic challenges faced by farmworkers during the COVID-19 pandemic (*9, 10*).

We initiated surveillance of SARS-CoV-2 infection among farmworkers in California’s Salinas Valley to monitor the epidemic. We have previously described impacts of the pandemic on economic well-being, mental health, and food insecurity within this population (*11*). Here we report on the prevalence of SARS-CoV-2 infection among individuals tested in farmworker-serving clinics from June to November, 2020, as well as symptoms and antibody responses within a subset of these farmworkers enrolled in a cross-sectional study.

## METHODS

### Study setting

The Salinas Valley is a 90-mile stretch of agricultural land within Monterey County, California; prominent farmed crops include leafy greens, berries, broccoli, artichokes, and wine grapes. The agricultural workforce comprises ∼50,000 resident farmworkers, with an additional 40,000 seasonal workers supporting the peak summer and fall seasons (*12*). The population is 75% Latino, and 30-60% of workers are believed to be undocumented (*13*). Severe overcrowding and household disrepair are common among farmworkers (*14*), with many living in multi-generational households (*15*) as well as labor camps, vehicles, and informal dwellings (*16*). Many farmworkers travel long distances to work, often in shared trucks or buses, and may work in close proximity to one another. These circumstances have led to concern about difficulty preventing SARS-CoV-2 transmission among farmworkers and in their communities (*17*).

We undertook this study in partnership with Clínica de Salud del Valle de Salinas (CSVS), a federally-qualified community and migrant health center in Monterey County. As the main healthcare provider for the region’s farmworkers and their families, CSVS operates a network of 12 comprehensive primary care centers serving 52,000 low-income, primarily Spanish-speaking patients.

### SARS-CoV-2 testing

Testing for SARS-CoV-2 infection at CSVS clinics began 15 June, 2020, and was offered to all individuals at clinics during weekday business hours. Medical personnel collected oropharyngeal specimens for detection of SARS-CoV-2 RNA via the qualitative Hologic (Marlborough, Massachusetts) Aptima nucleic acid transcription-mediated amplification (TMA) assay. All patients receiving care from CSVS for any reason were encouraged by their healthcare providers to receive SARS-CoV-2 testing, regardless of symptoms; testing was also made available to individuals who were not CSVS patients. No-cost testing for individuals without insurance was supported by funding from the Health Resources and Services Administration. Additionally, CSVS conducted outreach testing via mobile testing facilities at community sites including low-income and employer-provided housing, agricultural fields, homeless shelters, food banks, and CSVS-run health fairs where free SARS-CoV-2 testing was offered alongside seasonal influenza vaccination and food donations.

### Clinical surveillance study

We sought to compare the proportion of SARS-CoV-2 TMA tests yielding positive results among farmworkers against that among other adults tested by CSVS. As part of routine clinical intake, all patients ages ≥18 years were asked about employment. We considered farmworkers to include all persons engaged in work in agriculture, including but not limited to crop, nursery, and greenhouse laborers; agricultural equipment operators; workers in packing sheds and other food processing facilities; and farm and ranch animal workers and breeders.

### Cross-sectional study

#### Enrollment

To better understand the distribution, dynamics, and clinical profile of infection in this population, we invited farmworkers who were receiving a SARS-CoV-2 TMA test at CSVS to participate in a more in-depth cross-sectional study further entailing anti-SARS-CoV-2 antibody testing and a detailed questionnaire. To advertise the study, fliers were distributed in the community and to area growers and were hung in clinics describing the opportunity to receive free SARS-CoV-2 testing from CSVS and participate in the study. The study team was stationed at CSVS testing facilities and aimed to approach all patients receiving SARS-CoV-2 TMA tests to screen for study eligibility and invite participation in the cross-sectional study. When time allowed, study personnel called patients with scheduled SARS-CoV-2 testing appointments at CSVS on the day before their visit to advertise the study and screen for eligibility. Participants in an ongoing longitudinal study of farmworker families (*13*) and those living in housing for farmworkers were also invited to participate and to bring other farmworkers.

Eligible participants were non-pregnant adult farmworkers ages ≥18 years receiving SARS-CoV-2 TMA testing at CSVS who had conducted farm work ≤14 days before their testing date, who had not previously participated, and who spoke sufficient English or Spanish to give consent and complete study procedures. To accommodate the end of the growing season, from 5 October onward we enrolled individuals who had engaged in farm work any time since March 2020.

#### Study procedures

The study team obtained a blood sample by venipuncture and measured participants’ height and weight using large-print tape measures adhered to a post or wall and digital scales. Within 48 hours before (for pre-consented participants) or after the enrollment visit, and before SARS-CoV-2 testing results were available, the study team administered a 45-minute computer-guided questionnaire by telephone in Spanish or English. Questionnaire items addressed participant demographics, socio-economic status, symptoms since December 2019 and in the two weeks preceding enrollment, COVID-19 risk factors and exposures, and impacts of the pandemic on daily life and wellbeing (*11*). Following completion of all components of the study, the study team loaded a $50 incentive onto VISA gift cards handed out to participants at the enrollment visit.

Blood specimens were stored immediately at 4-7°C and centrifuged ≤48 hours after collection. Following separation, plasma aliquots were heat-inactivated at 56°C for 30 minutes and stored at –80°C, then used for assessment of immunoglobulin G (IgG) reactivity against the SARS-CoV-2 spike protein via in-house enzyme linked immunosorbent assays (ELISAs) (*18*). Briefly, recombinant full-length SARS-CoV-2 spike protein (John Pak, Chan Zuckerberg Biohub) was coated on Nunc Maxisorp ELISA plates (Themofisher) at 1.5µg/mL. Plates were blocked with 2.5% non-fat dry milk in 1X phosphate-buffered saline (PBS) for 2 hours at 37°C. Plates were then washed 3 times in 1X PBS. Plasma samples, diluted 1:100 in 1% non-fat dry milk in 1X PBS, were added to the plate in duplicate wells. After a 1-hour incubation at 37°C, plates were washed 5 times in 1X PBS + 0.05% Tween-20. Bound anti-spike IgG was detected using horseradish peroxidase-conjugated goat-anti-human IgG antibody (Fisher Scientific). The plate was developed using a 3,3′,5,5′-Tetramethylbenzidine (TMB) solution, and the reaction was stopped with 2M H2SO4 after 6 minutes. Prior assay validation was performed using convalescent (≥8 days post-onset) serum samples from 60 hospitalized, PCR-confirmed COVID-19 cases; 57 mild or subclinical PCR-confirmed COVID-19 cases; and 131 unexposed individuals (pre-2020 serum samples). Specimens were considered to be positive for anti-SARS-CoV-2 spike IgG if the ELISA optical density (OD) value was >0.096. This cutoff maximized area under the receiver operating characteristic curve, yielding 94.02% sensitivity and 98.47% specificity. All specimens were processed in duplicate; reflex testing was conducted if ≥1 OD measurement fell in the borderline range of 0.07-0.3 or if the coefficient of variation between replicates was ≥30% and ≥1 OD measure was ≥0.07. All specimens considered positive by the spike ELISA were confirmed by presence of IgG against the receptor binding domain (RBD) of SARS-CoV-2 spike protein (John Pak, Chan-Zuckerberg Biohub), using the protocol described above, substituting the coating antigen with RBD at 3ug/mL. Specimens were considered positive if RBD ELISA OD values were >0.205, determined via a similar validation process as described above for spike protein.

### Statistical analyses

#### Clinical surveillance study

We aggregated test results from all adult (≥18 years old) patients tested at CSVS from 15 June through 30 November, 2020. We examined the number with positive SARS-CoV-2 TMA test results by age, sex, and farmworker status. We also computed 2-week moving averages in the daily proportion of tests yielding positive results as well as estimates of the final proportion of tests yielding positive results by patient age, sex, and farmworker status. We used the Beta distribution to define 2.5% and 97.5% quantiles for the proportion positive.

#### Cross-sectional study

We measured associations of symptoms experienced in the last two weeks with current TMA-positive SARS-CoV-2 infection via the relative odds of reporting solicited symptoms (any symptoms, and each symptom individually) among individuals with positive or negative results; we obtained adjusted odds ratios (aORs) via logistic regression models that further accounted for age group (18-29, 30-39, 40-49, 50-59, or ≥60 years), sex, and recruitment venue (clinic-based or outreach). We used the same logistic regression framework to estimate aOR of experiencing each symptom since December 2019 or in the two weeks preceding enrollment with prior infection; for these analyses, anti-SARS-CoV-2 antibody OD measures (continuous) provided the exposure.

We next sought to estimate prevalence of current SARS-CoV-2 infection as well as seropositivity among farmworkers over the study period. Here we assumed individuals recruited via outreach testing represented the general farmworker population in the community in terms of symptoms, infection prevalence, and serostatus. We estimated stabilized sampling weights (*19*) to correct for differences in the population enrolled in the study over time. To generate weights for each recruitment period (16 July-31 August, 1-30 September, 1-31 October, or 1-30 November), we fit a multinomial logistic regression model which included all exposures listed in **Table 1**, the number of symptoms participants reported in the preceding two weeks, and the recruitment venue as predictors.

**Table 1:**
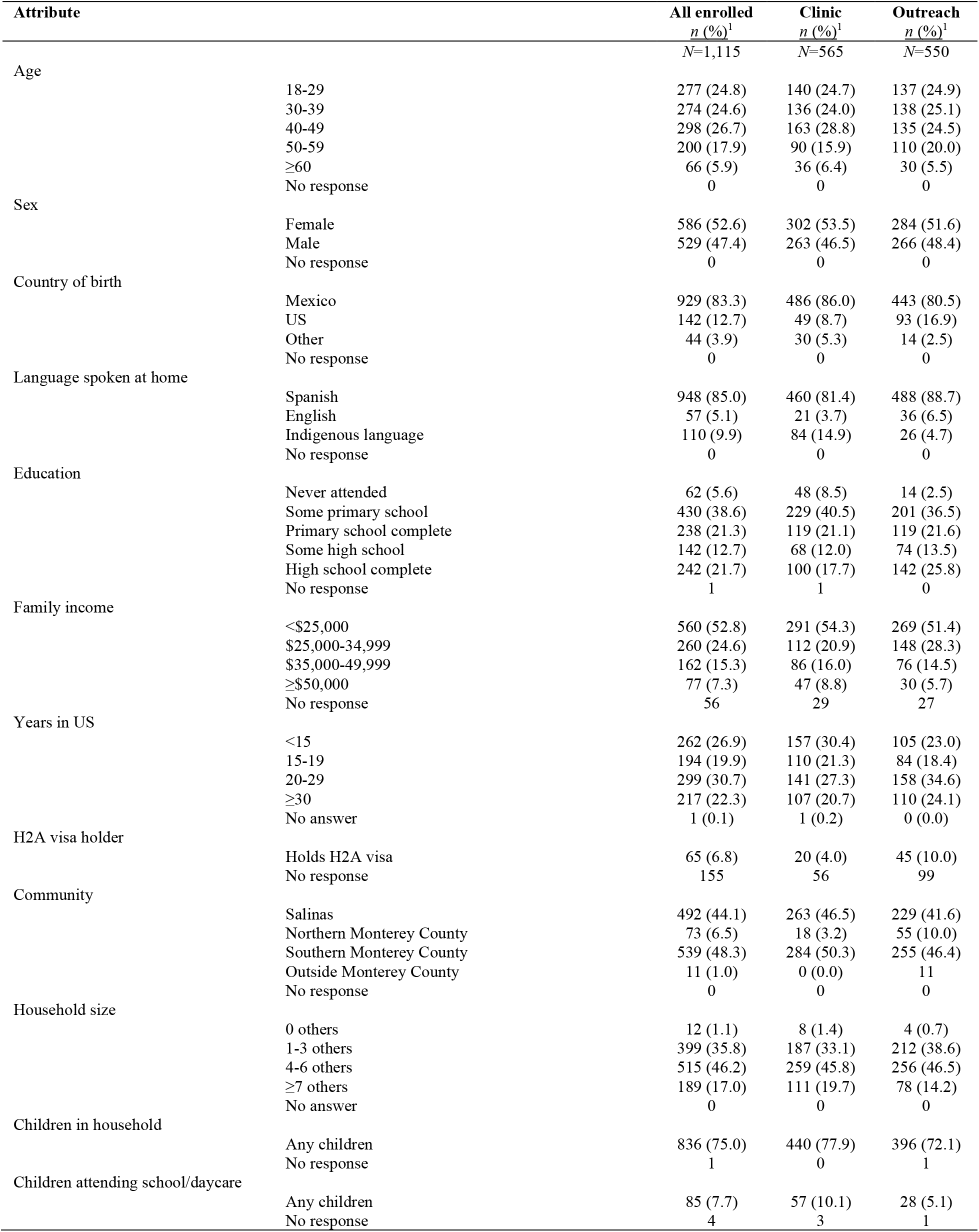
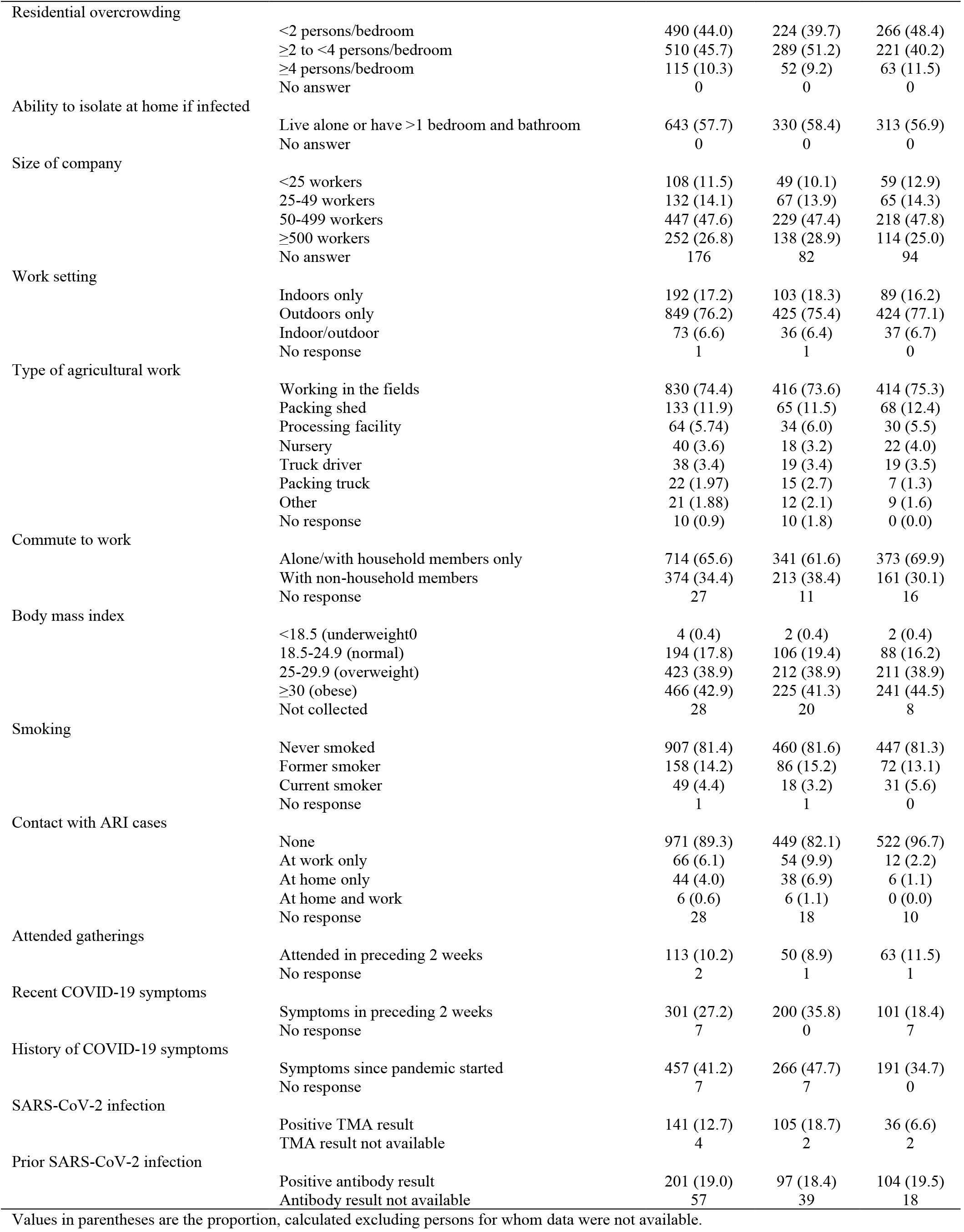
Characteristics of the study population, by recruitment setting, Monterey County farmworkers cross-sectional study, July 16 to November 30 2020, *N*=1115.

We estimated period-specific prevalence of SARS-CoV-2 infection and of seropositivity, accounting for inverse sampling weights, using a generalized linear model with a log-binomial link function. Models accounted for the four recruitment periods above, presence of any symptoms, and recruitment venue. We used the model parameter estimates to summarize period-specific prevalence of TMA-positive and seropositive status for individuals with and without symptoms, whom we would have expected to reach via community outreach. To account for missing data (1.1% of observations across all outcome and predictor variables), we sampled estimates from five independent iterations of the analysis carried out on multiply-imputed datasets. Analyses were conducted in R (version 4.0.3); we used the Amelia II package (*20*) for multiple imputation and fit the multinomial logistic model using the nnet package (*21*).

## RESULTS

### Clinical surveillance study

Between 15 June and 30 November, CSVS administered 14,169 SARS-CoV-2 TMA tests to adults, including 6,864 tests among farmworkers and 7,305 among other adults living in the same communities (**Figure 1A**). In total, 1,514 tests among farmworkers (22.1%) had positive results, as compared to 1,255 (17.2%) among other adults, corresponding to a 28.5% (95% confidence interval: 20.1-37.4%) higher probability of positive test results among farmworkers (**Figure 1B-C**). The test-positive fraction was similarly higher among men than among women both among farmworkers (23.7%, men vs. 20.5%, women; risk ratio: 1.16, 1.06-1.27) and non-farmworkers (21.7%, men vs. 18.8%, women; risk ratio: 1.15, 1.09-1.23). Point estimates of the test-positive fraction were consistent with equal or higher prevalence of infection among farmworkers across all age and sex strata (**Figure 1D-E**).

**Figure 1:**
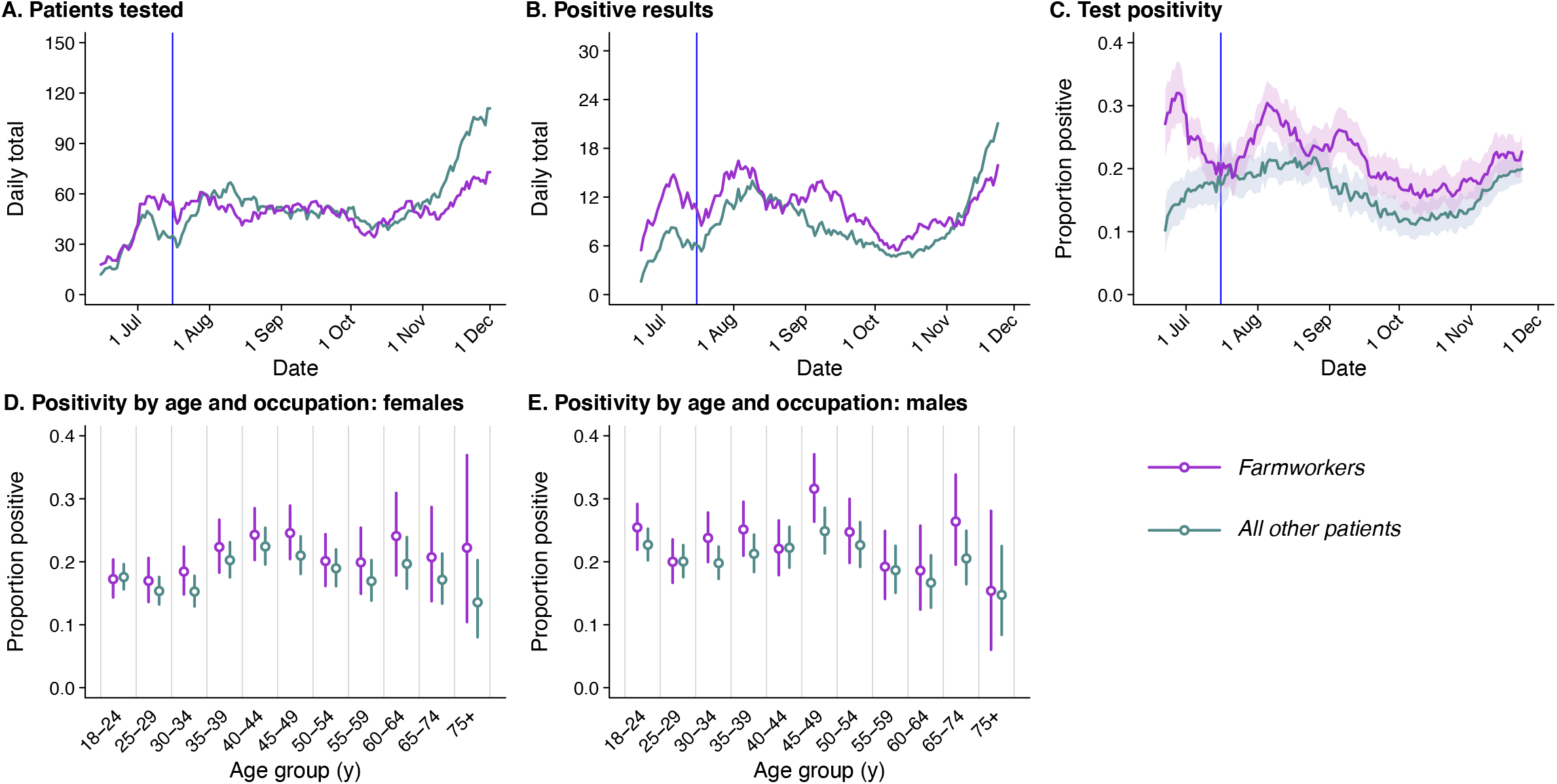
SARS-CoV-2 diagnoses at CSVS. We plot two-week moving averages of the (**A**) number of patients tested by CSVS; (**B**) the number of SARS-CoV-2 infection diagnoses; and (**C**) the proportion of tests yielding positive results, with shading for 95% confidence intervals. Below, we plot age- and sex-stratified test-positive fractions for (**D**) female and (**E**) male patients. Vertical lines in panels **A**-**C** indicate the date of initiation of the cross-sectional study (July 16, 2020).

Among farmworkers, multiple peaks in the proportion of TMA tests yielding positive results were evident, with the moving average of the test-positive fraction reaching 32.0% (27.2-37.0%) and 30.4% (27.0-34.0%) over the two-week periods surrounding 30 June and 14 August, respectively (**Figure 1C**). After declining from mid-September to early October, both the number of tests and the proportion yielding positive results increased through the remainder of the study period; from 10 October to 23 November, the two-week moving average of the number of tests conducted daily increased from 35.5 to 69.5 among farmworkers and from 38.7 to 104.5 among other adults; the proportion positive increased from 15.4% (12.2-18.8%) to 22.7% (20.0-25.5%) among farmworkers and from 12.1% (9.4-15.1%) to 19.9% (17.9-22.1%) among other adults. This increase in case volume among non-farmworker adults in November, without a commensurate rise among farmworkers, coincided with the annual migration of many Salinas Valley farmworkers to Yuma, Arizona and elsewhere (*22*).

### Cross-sectional study

Our cross-sectional study recruited 1,115 farmworkers, including 565 who were tested at clinics and 550 tested via outreach. SARS-CoV-2 TMA test results were obtained for 1,111 (99.6%) of these individuals, and ELISAs were conducted for 1,058 (94.9%; **Table 1**). Most of the farmworkers in this study were born in Mexico, spoke Spanish at home, had primary school-level education or less, earned <$25,000 per year, and worked in the fields; 36.3% lived in crowded housing. Most (81.8%) were overweight or obese, and only 4.4% were current smokers. In comparison to farmworkers recruited via outreach, farmworkers recruited at clinics had lower levels of educational attainment and a shorter length of stay in the US; a higher proportion spoke indigenous languages at home (14.9% vs. 4.7%) and reported contact with an individual experiencing respiratory symptoms (17.9% vs. 3.3%).

Overall, 27.2% of participants reported symptoms potentially related to COVID-19 in the last two weeks and 41.2% reported symptoms since the start of the pandemic. A higher proportion of farmworkers recruited via the clinic compared to those recruited via outreach reported at least one symptom potentially attributable to COVID-19 in either the two weeks before testing (35.8% vs. 18.4%; **Table 1**) or the period since December 2019 (47.7% vs. 34.7%). A total of 12.7% of all farmworkers tested TMA-positive for current SARS-CoV-2 infection, including 18.7% of farmworkers tested at clinics and 6.6% of those tested via outreach (**Table 1**). In contrast, 19.0% of farmworkers tested via ELISA were found to have antibody evidence of prior infection, with similar prevalence among those tested in the clinics (18.4%) and via outreach (19.4%).

Of all farmworkers who tested TMA-positive, 58.9% reported symptoms in the preceding two weeks, including 64.8% among those recruited from the clinic and 41.7% of those recruited via outreach (**Table 3**). Overall, 27.2% of those who had any potential COVID-19 symptoms in the two weeks prior to enrollment had current TMA-positive SARS-CoV-2 infection. Prevalence of current infection among farmworkers recruited in the clinic was 34.2% and 10.1% for those reporting any symptoms and no symptoms, respectively (**Table 2**). Among farmworkers recruited from outreach testing, current TMA-positive SARS-CoV-2 infection was detected in 14.9% and 4.7% of those reporting any symptoms and no symptoms, respectively. Following adjustment for age, sex, and recruitment setting, the adjusted odds ratio (aOR) of a TMA-positive SARS-CoV-2 test result was 4.17 (2.86-6.09) among farmworkers reporting any of the solicited symptoms of COVID-19 relative to those reporting no symptoms (**Table 3**). Symptoms most strongly associated with current SARS-CoV-2 infection included shortness of breath (aOR: 23.97; 8.15-73.27), loss of smell (aOR: 13.97; 6.29-31.41), loss of taste (aOR: 12.23; 5.67-26.59), and self-reported fever (aOR: 9.55; 5.30-17.22). Each of these symptoms, however, was reported by <25% of individuals with current SARS-CoV-2 infection. The most commonly reported symptoms among individuals testing positive, headache (32.6%) and myalgia (31.2%), were associated with 3.57-(2.34-5.46) and 6.15-(3.83-9.86) fold higher adjusted odds of SARS-CoV-2 RNA detection, respectively.

**Table 2:** Prevalence of SARS-CoV-2 infection by transcription-mediated amplification assay, symptoms, and recruitment setting.

| Symptom | Frequency of symptoms among all participants and test positivity among individuals with listed symptoms, *n* (%) |  |  |  |  |  |
| --- | --- | --- | --- | --- | --- | --- |
|  | *All participants* |  | *Clinic participants* |  | *Outreach participants* |  |
|  | Frequency<sup>1</sup> | Infected<sup>2</sup> | Frequency<sup>1</sup> | Infected<sup>2</sup> | Frequency<sup>1</sup> | Infected<sup>2</sup> |
|  | *N*=1,108 |  | *N*=558 |  | *N*=550 |  |
| Non-productive cough | 68 (6.1) | 31 (45.6) | 47 (8.4) | 26 (55.3) | 21 (3.8) | 5 (23.8) |
| Productive cough | 68 (6.1) | 25 (37.3) | 51 (9.1) | 21 (42.0) | 17 (3.1) | 4 (23.5) |
| Pain or pressure in the ears | 24 (2.2) | 10 (41.7) | 19 (3.4) | 10 (52.6) | 5 (0.9) | 0 (0.0) |
| Blocked nose | 62 (5.6) | 22 (36.1) | 50 (9.0) | 19 (38.8) | 12 (2.2) | 3 (25.0) |
| Runny nose | 78 (7.0) | 24 (31.2) | 56 (10.0) | 18 (32.7) | 22 (4.0) | 6 (27.3) |
| Sneezing | 95 (8.6) | 21 (22.3) | 61 (10.9) | 16 (26.7) | 34 (6.2) | 5 (14.7) |
| Watery eyes | 57 (5.1) | 14 (25.0) | 48 (8.6) | 14 (29.8) | 9 (1.6) | 0 (0.0) |
| Hoarseness | 49 (4.4) | 19 (38.8) | 42 (7.5) | 17 (40.5) | 7 (1.3) | 2 (28.6) |
| Self-reported fever<sup>3</sup> | 56 (5.1) | 33 (58.9) | 47 (8.4) | 29 (61.7) | 9 (1.6) | 4 (44.4) |
| Sweating | 48 (4.3) | 22 (45.8) | 40 (7.2) | 20 (50.0) | 8 (1.5) | 2 (25.0) |
| Chills | 74 (6.7) | 35 (47.3) | 63 (11.3) | 33 (52.4) | 11 (2.0) | 2 (18.2) |
| Headache | 147 (13.3) | 46 (31.5) | 100 (17.9) | 39 (39.4) | 47 (8.5) | 7 (14.9) |
| Tickle in throat | 49 (4.4) | 17 (34.7) | 36 (6.5) | 15 (41.7) | 13 (2.4) | 2 (15.4) |
| Sore throat | 103 (9.3) | 32 (31.1) | 78 (14.0) | 29 (37.2) | 25 (4.5) | 3 (12.0) |
| Myalgia | 97 (8.8) | 44 (45.8) | 79 (14.2) | 40 (51.3) | 18 (3.3) | 4 (22.2) |
| Chest pain | 26 (2.3) | 11 (42.3) | 21 (3.8) | 10 (47.6) | 5 (0.9) | 1 (20.0) |
| Sinus pain | 17 (1.5) | 7 (41.2) | 14 (2.5) | 7 (50.0) | 3 (0.5) | 0 (0.0) |
| Swollen glands | 18 (1.6) | 5 (27.8) | 11 (2.0) | 5 (45.5) | 7 (1.3) | 0 (0.0) |
| Loss of appetite | 38 (3.4) | 21 (55.3) | 32 (5.7) | 18 (56.2) | 6 (1.1) | 3 (50.0) |
| Difficulty breathing | 34 (3.1) | 18 (52.9) | 27 (4.8) | 16 (59.3) | 7 (1.3) | 2 (28.6) |
| Wheezing | 15 (1.4) | 6 (40.0) | 12 (2.2) | 6 (50.0) | 3 (0.5) | 0 (0.0) |
| Shortness of breath | 22 (2.0) | 18 (81.8) | 19 (3.4) | 16 (84.2) | 3 (0.5) | 2 (66.7) |
| Diarrhea | 40 (3.6) | 15 (37.5) | 33 (5.9) | 14 (42.4) | 7 (1.3) | 1 (14.3) |
| Nausea | 39 (3.5) | 13 (33.3) | 32 (5.7) | 13 (40.6) | 7 (1.3) | 0 (0.0) |
| Stomach pain | 47 (4.2) | 15 (31.9) | 34 (6.1) | 12 (35.3) | 13 (2.4) | 3 (23.1) |
| Trouble thinking | 18 (1.6) | 5 (27.8) | 10 (1.8) | 5 (50.0) | 8 (1.5) | 0 (0.0) |
| Fatigue | 94 (8.5) | 33 (35.5) | 70 (12.5) | 31 (44.9) | 24 (4.4) | 2 (8.3) |
| Loss of sense of taste | 33 (3.0) | 22 (66.7) | 26 (4.7) | 18 (69.2) | 7 (1.3) | 4 (57.1) |
| Loss of sense of smell | 32 (2.9) | 22 (68.8) | 25 (4.5) | 19 (76.0) | 7 (1.3) | 3 (42.9) |
| Pain or pressure in the eyes | 25 (2.3) | 6 (24.0) | 16 (2.9) | 6 (37.5) | 9 (1.6) | 0 (0.0) |
| No response | 7 |  | 7 |  | 0 |  |
| Any symptom | 301 (27.2) | 83 (27.7) | 200 (35.8) | 68 (34.2) | 101 (18.4) | 15 (14.9) |
| No symptoms | 807 (72.8) | 57 (7.1) | 358 (64.2) | 36 (10.1) | 449 (81.6) | 21 (4.7) |

<sup>1</sup>Proportions are computed among all tested
<sup>2</sup>Proportions indicate the prevalence of infection among those with the indicated symptom(s).
<sup>3</sup>Participants were not asked to verify whether they recorded their body temperature.

**Table 3:** Symptom prevalence and association with SARS-CoV-2 infection by transcription-mediated amplification assay.

| Symptom | Symptom frequency, *n* (%), by setting and infection status |  |  |  |  |  | Association with current infection |  |
| --- | --- | --- | --- | --- | --- | --- | --- | --- |
|  | *All participants* |  | *Clinic participants* |  | *Outreach participants* |  | OR (95% CI) | aOR (95% CI)<sup>1</sup> |
|  | TMA positive | TMA negative | TMA positive | TMA negative | TMA positive | TMA negative |  |  |
|  | *N*=141 | *N*=970 | *N*=105 | *N*=458 | *N*=36 | *N*=512 |  |  |
| Non-productive cough | 31 (22.1) | 37 (3.8) | 21 (24.8) | 21 (4.6) | 5 (13.9) | 16 (3.1) | 7.08 (4.21, 11.91) | 5.96 (3.51, 10.15) |
| Productive cough | 25 (17.9) | 42 (4.4) | 21 (20.0) | 29 (6.3) | 4 (11.1) | 13 (2.5) | 4.73 (2.78, 8.04) | 4.06 (2.34, 7.03) |
| Pain or pressure in the ears | 10 (7.1) | 14 (1.5) | 10 (9.5) | 9 (2.0) | 0 (0.0) | 5 (1.0) | 5.25 (2.28, 12.12) | 3.94 (1.67, 9.38) |
| Blocked nose | 22 (15.7) | 39 (4.0) | 19 (18.1) | 30 (6.6) | 3 (8.3) | 9 (1.8) | 4.48 (2.59, 7.76) | 3.43 (1.94, 6.04) |
| Runny nose | 24 (17.1) | 53 (3.1) | 18 (17.1) | 37 (8.1) | 6 (16.7) | 16 (3.1) | 3.54 (2.09, 5.92) | 3.02 (1.77, 5.14) |
| Sneezing | 21 (15.0) | 73 (7.6) | 16 (15.2) | 44 (9.6) | 5 (13.9) | 29 (5.7) | 2.11 (1.25, 3.56) | 1.99 (1.17, 3.39) |
| Watery eyes | 14 (10.0) | 42 (4.4) | 14 (13.3) | 33 (7.2) | 0 (0.0) | 9 (1.8) | 2.46 (1.30, 4.62) | 1.78 (0.94, 3.38) |
| Hoarseness | 19 (13.6) | 30 (3.1) | 17 (16.2) | 25 (5.5) | 2 (5.6) | 5 (1.0) | 4.81 (2.62, 8.83) | 3.34 (1.79, 6.22) |
| Self-reported fever<sup>2</sup> | 33 (23.6) | 23 (2.4) | 29 (27.6) | 18 (3.9) | 4 (11.1) | 5 (1.0) | 12.58 (7.15, 22.25) | 9.55 (5.30, 17.22) |
| Sweating | 22 (15.7) | 26 (2.6) | 20 (19.0) | 20 (4.4) | 2 (5.6) | 6 (1.2) | 6.62 (3.66, 12.04) | 4.83 (2.62, 8.95) |
| Chills | 35 (25.0) | 39 (4.0) | 33 (31.4) | 30 (6.6) | 2 (5.6) | 9 (1.8) | 7.94 (4.84, 13.07) | 5.95 (3.56, 9.92) |
| Headache | 46 (32.9) | 100 (10.4) | 39 (37.1) | 60 (13.1) | 7 (19.4) | 40 (7.8) | 4.22 (2.79, 6.34) | 3.57 (2.34, 5.46) |
| Tickle in throat | 17 (12.1) | 32 (3.3) | 15 (14.3) | 21 (4.6) | 2 (5.6) | 11 (2.1) | 4.07 (2.20, 7.54) | 3.27 (1.74, 6.16) |
| Sore throat | 32 (22.9) | 71 (7.4) | 29 (27.6) | 49 (10.7) | 3 (8.3) | 22 (4.3) | 3.75 (2.36, 5.95) | 2.98 (1.84, 4.82) |
| Myalgia | 44 (31.4) | 52 (5.4) | 40 (38.1) | 38 (8.3) | 4 (11.1) | 14 (2.7) | 8.00 (5.06, 12.54) | 6.15 (3.83, 9.86) |
| Chest pain | 11 (7.9) | 15 (1.6) | 10 (9.5) | 11 (2.4) | 1 (2.8) | 4 (0.8) | 5.12 (2.31, 11.38) | 3.81 (1.67, 8.64) |
| Sinus pain | 7 (5.0) | 10 (1.0) | 7 (6.7) | 7 (1.5) | 0 (0.0) | 3 (0.6) | 5.32 (1.98, 14.04) | 4.10 (1.51, 11.16) |
| Swollen glands | 5 (3.6) | 13 (1.3) | 5 (4.8) | 6 (1.3) | 0 (0.0) | 7 (1.4) | 2.71 (0.96, 7.72) | 2.55 (0.85, 7.62) |
| Loss of appetite | 21 (15.0) | 17 (1.8) | 18 (17.1) | 14 (3.1) | 3 (8.3) | 3 (0.6) | 9.47 (4.90, 18.42) | 7.17 (3.63, 14.17) |
| Difficulty breathing | 18 (12.9) | 16 (1.7) | 16 (15.2) | 11 (2.4) | 2 (5.6) | 5 (1.0) | 8.72 (4.35, 17.47) | 6.87 (3.36, 14.01) |
| Wheezing | 6 (4.3) | 9 (0.9) | 6 (5.7) | 6 (1.3) | 0 (0.0) | 3 (0.6) | 4.46 (1.58, 12.62) | 3.50 (1.18, 10.48) |
| Shortness of breath | 18 (12.9) | 4 (0.4) | 16 (15.2) | 3 (0.7) | 2 (5.6) | 1 (0.2) | 30.79 (10.71, 90.93) | 23.97 (8.15, 73.27) |
| Diarrhea | 15 (10.7) | 25 (2.6) | 14 (13.3) | 19 (4.1) | 1 (2.8) | 6 (1.2) | 4.42 (2.25, 8.58) | 3.38 (1.71, 6.75) |
| Nausea | 13 (9.3) | 26 (2.7) | 13 (12.4) | 19 (4.1) | 0 (0.0) | 7 (1.4) | 3.75 (1.87, 7.52) | 2.85 (1.42, 5.81) |
| Stomach pain | 15 (10.7) | 32 (3.3) | 12 (11.4) | 22 (4.8) | 3 (8.3) | 10 (2.0) | 3.44 (1.81, 6.56) | 2.72 (1.41, 5.27) |
| Trouble thinking | 5 (3.6) | 13 (1.3) | 5 (4.8) | 5 (1.1) | 0 (0.0) | 8 (1.6) | 2.77 (0.97, 7.75) | 2.57 (0.89, 7.46) |
| Fatigue | 33 (23.6) | 60 (6.2) | 31 (29.5) | 38 (8.3) | 2 (5.6) | 22 (4.3) | 4.61 (2.88, 7.40) | 3.74 (2.30, 6.07) |
| Loss of sense of taste | 22 (15.7) | 11 (1.1) | 18 (17.1) | 8 (1.7) | 4 (11.1) | 3 (0.6) | 15.07 (7.15, 31.82) | 12.23 (5.67, 26.59) |
| Loss of sense of smell | 22 (15.7) | 10 (1.0) | 19 (18.1) | 6 (1.3) | 3 (8.3) | 4 (0.8) | 17.28 (8.0, 37.79) | 13.97 (6.29, 31.41) |
| Pain or pressure in the eyes | 6 (4.3) | 19 (2.0) | 6 (5.7) | 10 (2.2) | 0 (0.0) | 9 (1.8) | 2.27 (0.89, 5.75) | 1.99 (0.76, 5.14) |
| No response | 1 | 6 | 1 | 6 | 0 | 0 | — | — |
| Any symptom | 83 (59.3) | 217 (22.5) | 68 (64.8) | 131 (28.6) | 15 (41.7) | 86 (16.8) | 4.96 (3.42, 7.16) | 4.17 (2.86, 6.09) |
| No symptoms | 57 (40.7) | 747 (77.5) | 37 (35.2) | 327 (71.4) | 21 (58.3) | 426 (85.2) | — | — |

<sup>1</sup>Logistic regression models adjusted for age group, sex, and recruitment venue.<sup>2</sup>Participants were not asked to verify whether they recorded their body temperature.

Individuals who recalled experiencing a blocked nose, sweating, chills, headache, a tickling sensation in the throat, a feeling of pain or pressure in the sinuses, loss of appetite, shortness of breath, fatigue, a loss of taste, a loss of smell since December 2019 had higher antibody reactivity, on average, than individuals who did not recall experiencing such symptoms (**Figure 2A**). We also identified higher antibody reactivity among individuals experiencing wheeze or loss of taste in the preceding two weeks, as well as suggestive associations of higher antibody measurements with chest pain and loss of smell in the preceding two weeks (**Figure 2B**). Quantitative antibody reactivity measures were not found to differ significantly among individuals who were or were not currently infected with SARS-CoV-2 (*p*=0.3), suggesting associations of antibody reactivity with recent symptoms were not attributable to current infection.

**Figure 2:**
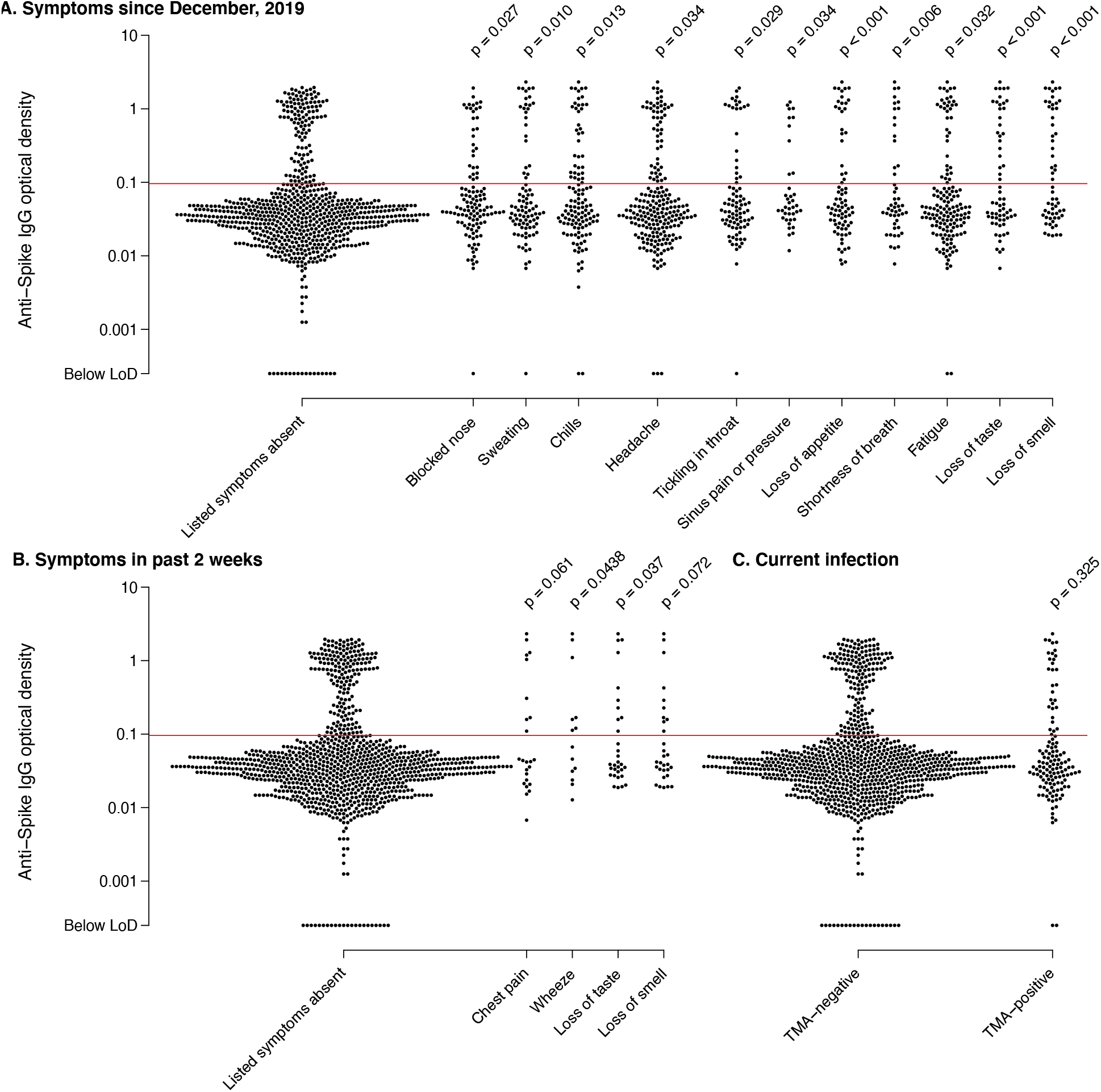
Anti-spike IgG antibody reactivity and association with recalled symptoms. We plot distributions of antibody reactivity (optical density, OD) among individuals who (**A**) reported experiencing or did not report experiencing various symptoms potentially associated with COVID-19 since December, 2019; (**B**) reported experiencing or did not report experiencing various symptoms in the two weeks before enrollment; and (**C**) had a positive or negative SARS-CoV-2 TMA test result at the enrollment visit; and. Reported *p*-values are measured in logistic regression models with the occurrence of each symptom as the outcome and antibody ELISA OD values (log-transformed) as predictors, with adjustment for age group and sex.

Last, we sought to estimate community prevalence of infection and seropositivity, reweighting the sample to adjust for differences in the population tested over time. We estimated the prevalence of current, TMA-positive SARS-CoV-2 infection was 5.6% (2.9-10.6%), 7.4% (4.4-12.4%), 4.5% (2.6-7.5%), and 8.0% (5.5-11.7%) over the periods of 16 July to 31 August, 1-30 September, 1-31 October, and 1-30 November, respectively (**Figure 3A**). These results closely tracked patterns in the proportion of tests yielding positive results among all farmworkers tested by CSVS (**Figure 1C**). Over this period, we estimated between 2.0% (0.9-4.4%) and 6.4% (4.0-10.2%) prevalence of current SARS-CoV-2 infection among asymptomatic persons in the community, and between 7.7% (3.7-15.8%) and 17.4% (10.4-29.3%) prevalence of current SARS-CoV-2 infection among individuals in the community experiencing ≥1 symptom. Estimated seroprevalence increased from 10.5% (6.0-18.4%) to 21.2% (16.6-27.4%) over the duration of the study, with similar results among symptomatic and asymptomatic individuals during each period (**Figure 3B**).

**Figure 3:**
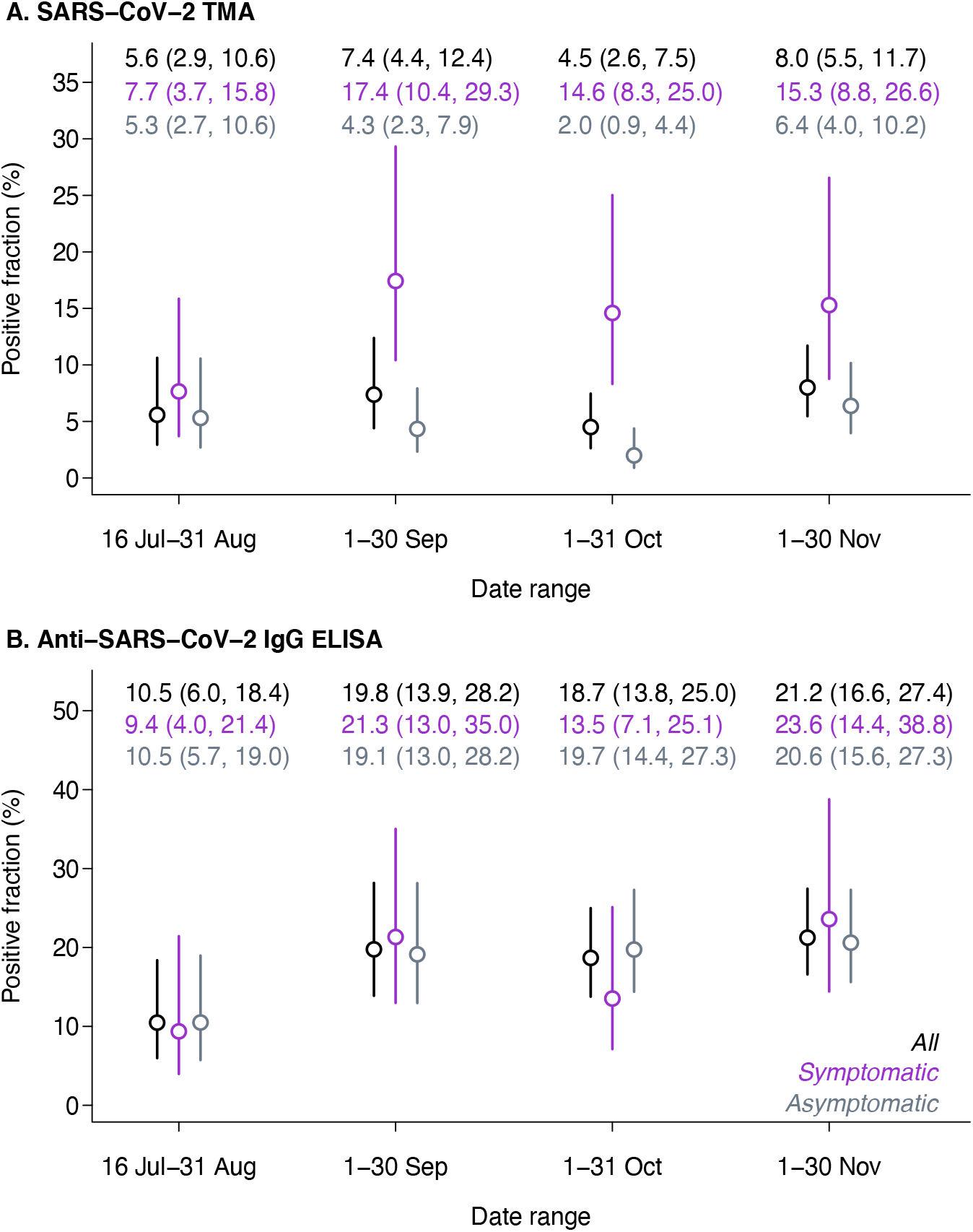
Community prevalence of SARS-CoV-2 positivity by TMA and seropositivity over time. We plot estimated prevalence of SARS-CoV-2 infection and seropositivity in samples reweighted to correct for differences in the population seeking testing over the course of the study. Lines delineate 95% confidence intervals around point estimates.

## DISCUSSION

Among all adults tested for SARS-CoV-2 infection by clinics serving the Monterey County farmworker population, test positivity was 28% higher for farmworkers than for non-farmworkers from the same communities. In comparison to the 6.1% overall test-positive fraction observed in Monterey County over the same period (*23*), test-positivity was 362% higher among farmworkers tested by CSVS. Within the cross-sectional study subpopulation, we identified sustained, high prevalence of infection, with TMA-positive results among 6.6% of individuals tested in the community and 18.7% of those tested in clinics. We estimated that roughly 10% of the farmworker population became infected over a 3-month period during the study, yielding ∼21% seroprevalence by November, 2020. This seroprevalence is well above the 5% seroprevalence noted among California adults in a large-scale assessment of blood specimens submitted for routine clinical screening or clinical management in September (*24*). A previous study in San Francisco likewise identified elevated infection risk in an urban, low-income and predominantly Latino population, with 6.0% prevalence of current infection among frontline workers and. 7.7% seroprevalence as of late April (*25*). Our findings demonstrate high infection risk among farmworkers during the ongoing pandemic.

We identified a diverse array of symptoms, including gastrointestinal and other non-respiratory symptoms, associated with SARS-CoV-2 infection. However, 41% of individuals found to be TMA-positive for current SARS-CoV-2 infection in our study did not report experiencing any symptoms. This figure is likely higher than the true fraction of asymptomatic infections, as individuals could have been pre-symptomatic at the time of their interview (*26*), the estimated 2-6% prevalence of infection among individuals without symptoms in the community suggests substantial risk of exposure to clinically-inapparent cases. Guidance issued for growers to screen farmworkers for fever or other COVID-19 symptoms is thus likely inadequate to prevent workplace infections (*27*). We also identified associations of higher antibody reactivity with current symptoms including loss of taste and smell, chest pain, and wheeze. Participants in our study likely experienced these symptoms in a persisting manner beyond the acute infectious stage, as seroconversion typically occurs 8-14 days following initial symptoms (*28*). While the clinical profile of “long COVID” remains to be fully clarified, these same symptoms have been identified as prominent complaints in prior studies along with fatigue, joint pain, and headache (*29*–*31*).

Our study has limitations. We cannot verify how well our sample represents the farmworker population, many of whom are “hidden” from population statistical measures (*32*). As we excluded individuals who did not speak Spanish or English sufficiently well to participate in the cross-sectional study, our study likely under-represents indigenous populations (estimated at 13% of Salinas Valley farmworkers (*12*)). Roughly half of our cross-sectional study participants were enrolled in clinic-based testing, among whom infection prevalence could be expected to be higher; nonetheless, many of these individuals did not experience COVID-19 symptoms, and our statistical framework accounted for differences between clinic-based and outreach samples. Last, waning antibody titers from infections acquired early in the pandemic may have contributed to under-estimation of seroprevalence, in particular for individuals who experienced mild or asymptomatic infection (*33*).

While Phase 1 vaccination programs have prioritized residents of long-term care facilities and healthcare workers, at the recommendation of the Advisory Committee on Immunization Practices (*34*), prioritization of differing essential workforce groups among Phase 2 recipients will be determined by states. Our study demonstrates high risk of SARS-CoV-2 infection and both acute and persisting symptoms of COVID-19 among farmworkers in California’s Salinas Valley, underscoring the urgency of preventive interventions for this population.

## Data Availability

Direct data requests to the corresponding author at

## ACKNOWLEDGMENTS

This study received financial support from the Innovative Genomics Institute on the UC Berkeley Campus. JAL discloses receipt of grants and fees from Pfizer unrelated to this study. All other authors declare no conflicts of interest.

The study was reviewed and approved by the Committee for Protection of Human Subjects at UC Berkeley.

Members of the CHAMACOS-Project-19 study team include: Jose Camacho, Gardenia Casillas, Celeste Castro, Cynthia Chang, Lupe Flores, Lizari Garcia, Madison J. de Vere, Maria Reina Garcia, Terry Gomez, Carly Hyland, Daniel Lampert, Aaron McDowell-Sanchez, Dominic Pina Montes, Jacqueline Montoya, Lilibeth Nunez, Juanita “Liz” Orozco, Marbel Orozco, Nargis Rezai, Maria T. Rodriquez, Monica Romero, Hina Sheth, Jon Yoshiyama, and Litzi Zepeda

## AUTHOR BIO

Joseph Lewnard is an Assistant Professor of Epidemiology at the School of Public Health, University of California, Berkeley, and studies infectious disease transmission dynamics and control. Ana M. Mora is an Assistant Researcher at the University of California, Berkeley and an Associate Professor at Universidad Nacional, Heredia, Costa Rica, and primarily studies of the health effects of exposures to environmental toxicants.

## Notes

### Competing Interest Statement

JAL has received grants and fees from Pfizer, Inc. unrelated to the submitted work.

